# Testing and Vaccination to Reduce the Impact of COVID-19 in Nursing Homes: An Agent-Based Approach

**DOI:** 10.1101/2021.03.22.21254125

**Authors:** Jose Pablo Gómez-Vázquez, Yury García, Alec J. Schmidt, Beatriz Martínez-López, Miriam Nuño

**Affiliations:** Center for Animal Disease Modeling and Surveillance, University of California Davis, CA, USA; Department of Public Health Sciences, University of California Davis, CA, USA

**Keywords:** Nursing Homes, Testing, Vaccine, COVID-19, Agent-Based Model

## Abstract

**Background:** Efforts to protect residents in nursing homes involve non-pharmaceutical interventions, testing, and vaccine. We sought to quantify the effect of testing and vaccine strategies on the attack rate, length of the epidemic, and hospitalization.

**Methods:** We developed an agent-based model to simulate the dynamics of SARS-CoV-2 transmission in a nursing home with resident and staff agents. Interactions between 172 residents and 170 staff were assumed based on data from a nursing home in Los Angeles, CA. We simulated scenarios assuming different levels of non-pharmaceutical interventions, testing frequencies, and vaccine efficacy to block transmission.

**Results:** Under the hypothetical scenario of widespread SARS-CoV-2 in the community, 3-day testing frequency minimized the attack rate and the time to eradicate an outbreak. Prioritization of vaccine among staff or staff and residents minimized the cumulative number of infections and hospitalization, particularly in the scenario of high probability of an introduction. Reducing the probability of a virus introduction reduced the demand on testing and vaccine to reduce infections and hospitalizations.

**Conclusions:** Improving frequency of testing from 7-days to 3-days minimized the number of infections and hospitalizations, despite widespread community transmission. Vaccine prioritization of staff provides the best protection strategy, despite high risk of a virus introduction.

## Introduction

COVID-19 has highlighted many inadequacies in the American healthcare system. Elderly and frail residents of long-term care facilities (LTCFs) have experienced a disproportionate burden of infection and death. Approximately 5% of all US cases have occurred in LTCFs, yet deaths related to COVID-19 in these facilities account for 34% of all US deaths as of February 12, 2021, according to the New York Times [1]. Nationwide, there are about 44,736 LTCFs in the United States, 15,116 of which are nursing homes. Together these facilities encompass more than 1.2 million staff and 2.1 million residents based on 2015-2016 estimates [2].

Guidance on the prevention and mitigation of COVID-19 in LTCFs was offered by many oversight groups, including the Centers for Disease Control and Prevention (CDC) and the Center for Medicare and Medicaid Services (CMS). Substantial numbers of transmission events from symptom-free individuals made it clear that universal testing, regardless of symptoms, was a critical component of a robust prevention program [3–5]. Testing frequency was widely debated, as LTCFs had to balance the obvious need with the high cost and low availability of testing, especially early in the pandemic [5–7]. Vaccines provide an incredible tool for preventing COVID-19 outbreaks in LTCFs, but they are not a magic solution, nor will they be distributed into an environment that is wholly prepared to implement new protective measures.

Nursing home residents are a priority group for vaccination, as are health care workers. The CDC launched the Pharmacy Partnerships for Long-Term Care Program in an effort to provide on-site vaccination to residents and staff members in LTCFs [8]. Despite deployment of vaccines in LTCFs appearing successful thus far, there is a growing concern that insufficient levels of vaccine coverage will be reached. As of the end of January 2021, median first dose rates among LTCF residents is 77.8%, but only a median of 37.5% of staff have received at least their first dose [9]. It is unclear at this time whether the lower vaccination rates among staff is a result of prioritization of residents, lack of recording alternative sources of vaccination, or staff choice; however, a survey of nursing home staff conducted in the state of Indiana (November 2020) found that 45% of respondents were willing to receive a COVID-19 vaccine immediately once available, and an additional 24% would consider it in the future [10]. While visitors are disallowed and residents only interact directly with a small number of other people, staff are the primary vector for viral introduction [11, 12]; therefore, low rates of vaccine uptake among staff should be of great concern from the perspective of preventing an outbreak. Additionally, there is limited evidence about the ability of vaccines to reduce asymptomatic transmission. Preliminary data from the UK suggests a 49.3% reduction in infections from an asymptomatic source [13]. Recent evidence of the circulation of more transmissible SARS-CoV-2 variants also raises concerns about the course of this pandemic, particularly as less than 7% of the US population have received the full vaccine dosage [14].

Given the continued challenge of implementing robust protective measures in LTCFs, the bevy of unknowns around vaccine deployment, the uncertainty involved with new circulating strains, and the impending easing of co-recreation and visitor restrictions as states ease recommendations, we sought to quantify the effect of testing rates and differing vaccination strategies on morbidity and mortality in a long term care setting, using a nursing home in Los Angeles, CA as the foundation for an agent-based model (ABM). Our study assumes the continued presence of non-pharmaceutical interventions (NPIs) such as mask mandates for staff and universal testing, and varies the risk of introduction by staff. The end results is a model that can be adapted/modified to study the effects of these interventions in varied nursing home settings. Such modelling approaches can provide valuable insight into the design and deployment of combined vaccine and surveillance interventions before primary prospective research can be implemented [12].

## Methods

### Model structure

We developed a stochastic agent-based model to simulate the spread of SARS-CoV-2 in an LTCF, based on the floor plan and occupancy of a nursing home in Los Angeles County, California with 172 residents and 170 staff [Figure 1]. The simplified floor map shows the location of bedrooms with a capacity of 3 residents, 5 quarantine rooms reserved for residents with frequent outside traffic and/or capacity to quarantine exposed residents, recreation areas which are currently off limits to resident and staff interactions, and rooms for staff.

**Figure 1:**
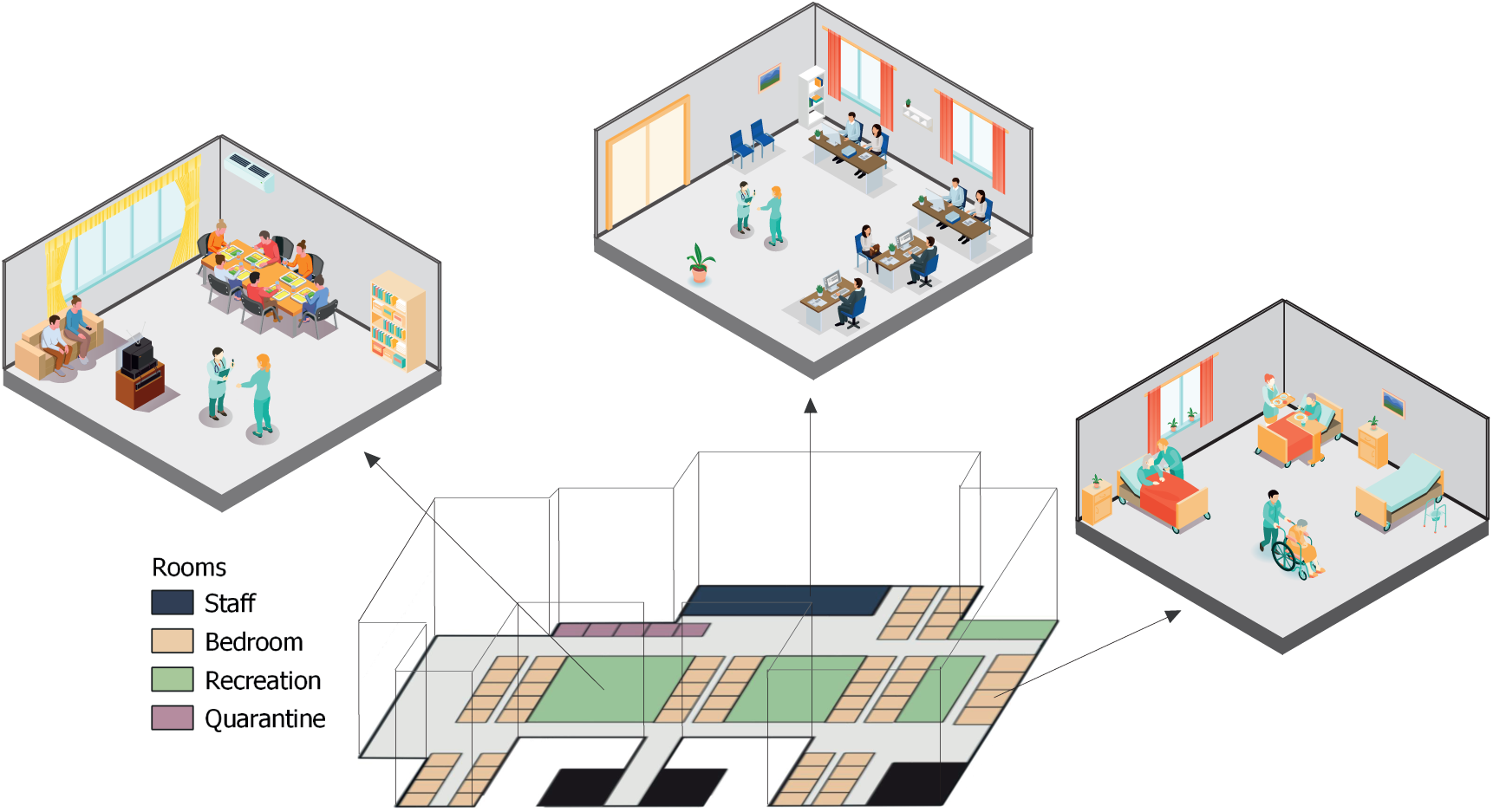
Case study of a nursing home in Los Angeles, CA.

Agents in the model include residents and staff, with the natural history of COVID-19 captured through seven epidemiological classes (Figure S1). The model assumes that residents are not replaced with new susceptible agents, and staff with confirmed exposure to the virus are replaced by new staff confirmed negative for SARS-CoV-2 during the period of simulation. Recovered people gain immunity to reinfection lasting 120 days, and the latency period is sampled from a logarithmic normal distribution [15]. Parameters and sources are described in Table 1.

**Table 1:** Parameter Descriptions, Baseline Values, and References.

| Description | Baseline Value | Ref. |
| --- | --- | --- |
| Average time a person remains in the non-infectious latency state ($\alpha$) | *lognormal*(7, 3)<sup>b</sup> | [15] |
| Proportion of asymptomatic people ($f$) | 0.40 | [16] |
| Average recovery time ($\gamma_1$) | 15 days | [17] |
| Proportion of hospitalized people ($\sigma_1$) | 0.23 | [18, 19] |
| Median number of days from symptom onset to hospitalization ($\gamma_2$) | 4 (1, 9) days | [20] |
| Median number of days of hospitalization ($\gamma_3$) | 6 (3, 10) days | [20] |
| Percent that die among those hospitalized. ($\sigma_2$) | 11.8% | [21] |
| Shedding probability | 0.38 | [a] |
| Infection probability | 0.38 | [a] |
| Introduction probability | 0.1 | [a] |
| **Assumptions for the scenarios** |  |  |
| Percentage of staff using PPE | 90% | [a] |
| Percentage of residents using PPE | 75% | [a] |
| PPE Effect ($OR_{pi}$) | 0.1467 | [a] [22] |
| Test detection probability | 80% | [a] |
| Percentage of Staff tested | 90% | [a] |
| Percentage of Resident tested | 33.3% | [a] |
| Frequency of testing | *Weekly* | [a] |
| Vaccine effect ($OR_v$) | 0.0493 | [a] [23] |
| Vaccine immunity duration | 120 days | [a] |
| **Distribution of the staff agent characteristics** |  |  |
| CN Contacts per hour | *Multinom* $\sim (X_0 = 0.7, X_1 = 0.3)$ | |
| RN Contacts per hour | *Multinom* $\sim (X_0 = 0.25, X_1 = 0.75)$ | |
| LPN Contacts per hour | *Multinom* $\sim (X_0 = 0.15, X_2 = 0.2, X_3 = 0.25, X_4 = 0.2, X_5 = 0.2)$ | |
| Work Schedule | *Multinom* $\sim (X_{morning} = 0.4, X_{night} = 0.2)$ | |
| Staff type | *Multinom* $\sim (X_{CN} = 0.6, X_{RN} = 0.15, X_{LPN} = 0.15)$ | |

<sup>a</sup>Explored via sensitivity analysis, <sup>b</sup>fitted to a distribution from data and truncated to a range of plausible values

### Disease dynamics

The transmission of the virus is based on the probability of contact between susceptible people and those who are in presymptomatic, asymptomatic, or symptomatic states. Each infectious state has the same probability of infecting others on contact. Due to default preventive testing and isolation measures, only *I*_*a*_ and *I*_*s*_ agents that have not been detected and isolated may contribute to new infections. A newly-infected individual enters a latency period sampled from a lognormal distribution with a mean of 7 days [15]. After that time, 40% of people remain asymptomatic [16] until recovery. For those who develop symptoms, 23% [18, 19] require hospitalization. The average number of days from the onset of symptoms to hospitalization is 4 days and a person stays in the hospital for an average of 6 days [20]. Mortality rate was set at 11.8% [21] for hospitalized agents. The average recovery time for asymptomatic agents or those who never required hospitalization is 15 days [17], during which they remain infectious. We assumed that recovery from a primary infection provided adequate immunity for the remainder of the simulation.

### Staff and resident interactions

Agents in the model include residents and staff only, consistent with the full visitor restrictions. Three residents are assigned to a single room. Five rooms are designated for quarantine/isolation of infected patients or for residents who require outside specialty care, such as dialysis. Residents only interact with two other residents in the same room and with staff, who can be one of three types: Certified Nursing Assistant (CNA), Registered Nurse (RN), and Licensed Practical Nurse (LPN). Since meals are taken in rooms and use of communal space is restricted, residents do not currently interact with residents outside assigned rooms.

Each type of staff has different contact patterns with residents throughout the day. These contact rates are operationalized as contact probabilities defined from a multinomial distribution where each hour a CNA has a 0.7 chance to have 0 contacts and 0.3 chance to have 1 contact with a resident, a LPN has a 0.15 chance of having 0 contacts, 0.2 of two constants, 0.25 chance of having 3 contacts, and so on (Table 1). Contact probability parameters were estimated from staff hour-per-resident-day (HRD) data from the CMS Nursing Home Compare data set. We assumed no difference in probability of viral introduction by staff type. Staff are assigned to one of three different work schedules: 40% work in the morning (7am-3pm), 40% in the afternoon (3-11 pm), and 20% work overnight. They spend on average 8 hours inside the nursing home and the rest of the time in the community. Both scheduled time and type of staff are sampled from a multinomial distribution to reflect the distribution in our reference nursing home (Table 1).

### COVID-19 transmission in the community

Though there is large variability on the impact of COVID-19 in these facilities, tied to historic variability in testing capacity and PPE availability and adherence, the most immediate risk of a COVID-19 outbreak in a nursing home is the level of community transmission of SARS-CoV-2. Since we assumed that visitors are disallowed completely, residents’ risk for primary exposure is contact through staff who acquired an infection from the wider community. A critical factor that our model aimed to study was to assess the impact of the probability of viral introduction from the community on the predicted size of internal outbreaks. Each scenario we investigated was simulated across three different probabilities of a staff member introducing the infection: low (5% per day), medium (10% per day), and high (15% per day). These are expressed as ‘introduction probability’, which is set to 0.1 for the baseline scenario 1.

### Interventions

We parameterized interventions with variable impacts on the transmission of SARS-CoV-2: PPE use and misuse, regular diagnostic testing, and vaccinations. We considered scenarios where staff were tested every 7 days (baseline), 5 days, and 3 days. Testing of residents in all scenarios assume that one resident per room is tested weekly, systematically cycling through the each resident every three weeks. Reduction in transmission probability from PPE use and vaccination were applied by modifying the shedding and infection probability parameters (Table *S*1). Vaccine efficacy was translated into odds ratios of infection given exposure from the Pfizer and Moderna phase 3 clinical trial results. For brand- and age-agnostic scenarios, including the baseline scenario, the crude overall odds ratio was set to 0.0493. In scenarios where vaccine brand and recipient age were taken into account, the efficacy of the Moderna vaccine after the second dose was 95.6% (OR 0.0441) for individuals under 65 years old and 86.4% (OR 0.1357) for 65 and older [23]. The efficacy of the Pzifer vaccine for individuals under 65 was roughly equivalent to Moderna (OR 0.434), but was 94.7% (OR 0.0619 for individuals 65 years and older [24]. For ease of implementation, residents were considered 65 and older, and staff were considered under 65. The vaccine odds ratio has a direct impact on transmission probabilities and reflects the upper bounds for vaccine efficacy according to Equation 1. Let *p*_*t*_ be the probability of a transmission event:

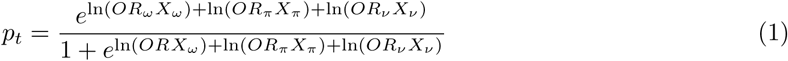

where the odds ratio *ω* (*OR*_*ω*_) represents the global baseline transmission probability of all agents, the odds ratio *π* (*OR*_*π*_) represents the transmission reduction from the presence or absence of PPE, and the odds ratio *v* (*OR*_*v*_) corresponds to the effect of vaccine status on transmission. Probability _*t*_ is computed for all agents at each time step in order to reflect different probabilities of transmission based on the interventions each individual received. For scenarios where a vaccine was implemented, we specified the proportion of residents and staff that received a vaccine and a fixed time interval of 21 days between the first and second dose, with a 60% efficacy after the first dose but before the second.

### Model Scenarios

The baseline scenario assumed current CDC infection prevention and control recommendations for nursing homes, including visitor restrictions, daily symptom screening of residents and staff, use of face masks, and weekly testing of staff. We incorporated weekly cyclic testing of one of three residents per room, with alternating residents being tested each week. When a resident tested positive, they were isolated and the other residents from the same room were tested. Staff who tested positive were ‘isolated’ (removed from the simulation, as if on paid leave) and replaced with new staff who tested negative. Parameters assumed for the baseline scenario are described on Table 1. We simulated a set of scenarios based on staff testing frequency, prioritization of residents or staff for vaccination, and vaccine brand. In each scenario we systematically changed one of these approaches while holding the others at baseline values. Outcomes were estimates of the length of the outbreak, total number of infections (and attack rate), hospitalizations, and deaths across three rates of community transmission (low, medium, and high).

### Model implementation

The model was implemented in GAMA 1.8.1 [25]. Code for reproducing this study is available at https://github.com/jpablo91/NH_COVID. Each scenario was simulated 200 times, and the median and 95% confidence intervals for each outcome were reported. For each set of simulations, we used the same seed to conduct sensitivity analysis and make comparisons between scenarios. The model was calibrated with data on confirmed COVID-19 cases reported between May 24, 2020 and February 14, 2021 in California nursing homes with similar resident census, extracted from the Centers for Medicare Medicaid Services (CMS) [26]. We considered a good fit to be have high *R*^2^ and Pearson’s *R* estimates between the observed and model-predicted cumulative number of confirmed cases.

## Results

### Baseline Scenario

In the baseline scenario we assumed PPE mandates, weekly testing, and no vaccination. Baseline attack rate was 0.17 (95% CI: 0.01, 0.39) and a median time to eradication of 28 days (Table 2, Figure 2). With the implementation of the vaccine and under the scenario of a high probability of introduction, the attack rate goes down to 0.02 and the time to the eradication of an outbreak was 14 days. (Figure 2, Suppl. Table 2).

**Table 2:** Results from the sensitivity analysis summarized by the median and 95% confidence intervals for the various simulations considered.

| Target Parameter | Value used | Days to Eradication | Attack Rate | Total Infected | Infected residents | Infected staff | Hospitalizations |
| --- | --- | --- | --- | --- | --- | --- | --- |
| Baseline <sup>a</sup> |  | 28 (7, 70) | 0.17 (0.01, 0.39) | 59 (2, 134) | 32 (0, 82) | 23 (1, 53) | 5 (0, 15) |
| Transmission probability |  |  |  |  |  |  |  |
| Low transmission virus<sup>a</sup> | 0.34 | 21 (7, 63) | 0.03 (0, 0.27) | 12 (1, 93) | 5 (0, 58) | 8 (1, 38) | 1 (0, 11) |
| High transmission virus<sup>a</sup> | 0.42 | 28 (7, 63) | 0.27 (0.01, 0.47) | 93 (2, 163) | 56 (0, 100) | 37 (1, 65) | 9 (0, 18) |
| Introduction probability |  |  |  |  |  |  |  |
| Low introduction probability<sup>a</sup> | 0.05 | 14 (7, 49) | 0.02 (0, 0.32) | 7 (0, 110) | 3 (0, 72) | 5 (0, 42) | 1 (0, 14) |
| High introduction probability<sup>a</sup> | 0.15 | 28 (9, 77) | 0.21 (0.01, 0.42) | 72 (3, 143) | 39 (0, 84) | 30 (2, 58) | 6 (0, 15) |
| Detection probability |  |  |  |  |  |  |  |
| Low detection probability (test)<sup>a</sup> | 0.7 | 28 (7, 70) | 0.2 (0, 0.47) | 68 (1, 161) | 38 (0, 97) | 30 (1, 62) | 5 (0, 21) |
| High detection probability (test)<sup>a</sup> | 0.9 | 21 (7, 70) | 0.09 (0, 0.31) | 32 (1, 108) | 17 (0, 67) | 16 (1, 44) | 3 (0, 12) |
| PPE effect |  |  |  |  |  |  |  |
| High effect PPE<sup>a</sup> | 0.07 | 15 (7, 56) | 0.02 (0, 0.2) | 7 (1, 69) | 2 (0, 40) | 5 (1, 31) | 1 (0, 7) |
| Low effect PPE<sup>a</sup> | 0.34 | 21 (14, 42) | 0.66 (0.39, 0.85) | 228 (134, 294) | 140 (79, 182) | 88 (49, 114) | 19 (6, 30) |
| Testing frequency |  |  |  |  |  |  |  |
| Testing frequency, 5-days<sup>a</sup> | 5-days | 15 (5, 49) | 0.03 (0, 0.22) | 10 (1, 74) | 3 (0, 44) | 7 (1, 30) | 1 (0, 9) |
| Testing Frequency, 3-days<sup>a</sup> | 3-days | 12 (3, 36) | 0.01 (0, 0.15) | 5 (1, 51) | 1 (0, 34) | 4 (1, 21) | 1 (0, 5) |
| Vaccine effect |  |  |  |  |  |  |  |
| Similar age-specific vaccine efficacy | 0.04 | 14 (7, 35) | 0.01 (0, 0.05) | 4 (1, 17) | 0 (0, 9) | 3 (0, 11) | 0 (0, 2) |
| Different age-specific vaccine efficacy [Pfizer] | 0.06<sup>b</sup>, 0.04<sup>c</sup> | 13 (7, 38) | 0.01 (0, 0.06) | 4 (1, 20) | 0 (0, 9) | 3 (0, 11) | 0 (0, 2) |
| Different age-specific vaccine efficacy [Moderna] | 0.13<sup>b</sup>, 0.04<sup>c</sup> | 14 (7, 41) | 0.01 (0, 0.07) | 5 (1, 24) | 1 (0, 14) | 3 (1, 14) | 0 (0, 3) |

<sup>a</sup>No vaccination assumed, <sup>b</sup> Vaccine assumed for resident agents, <sup>c</sup> Vaccine assumed for staff agents

**Figure 2:**
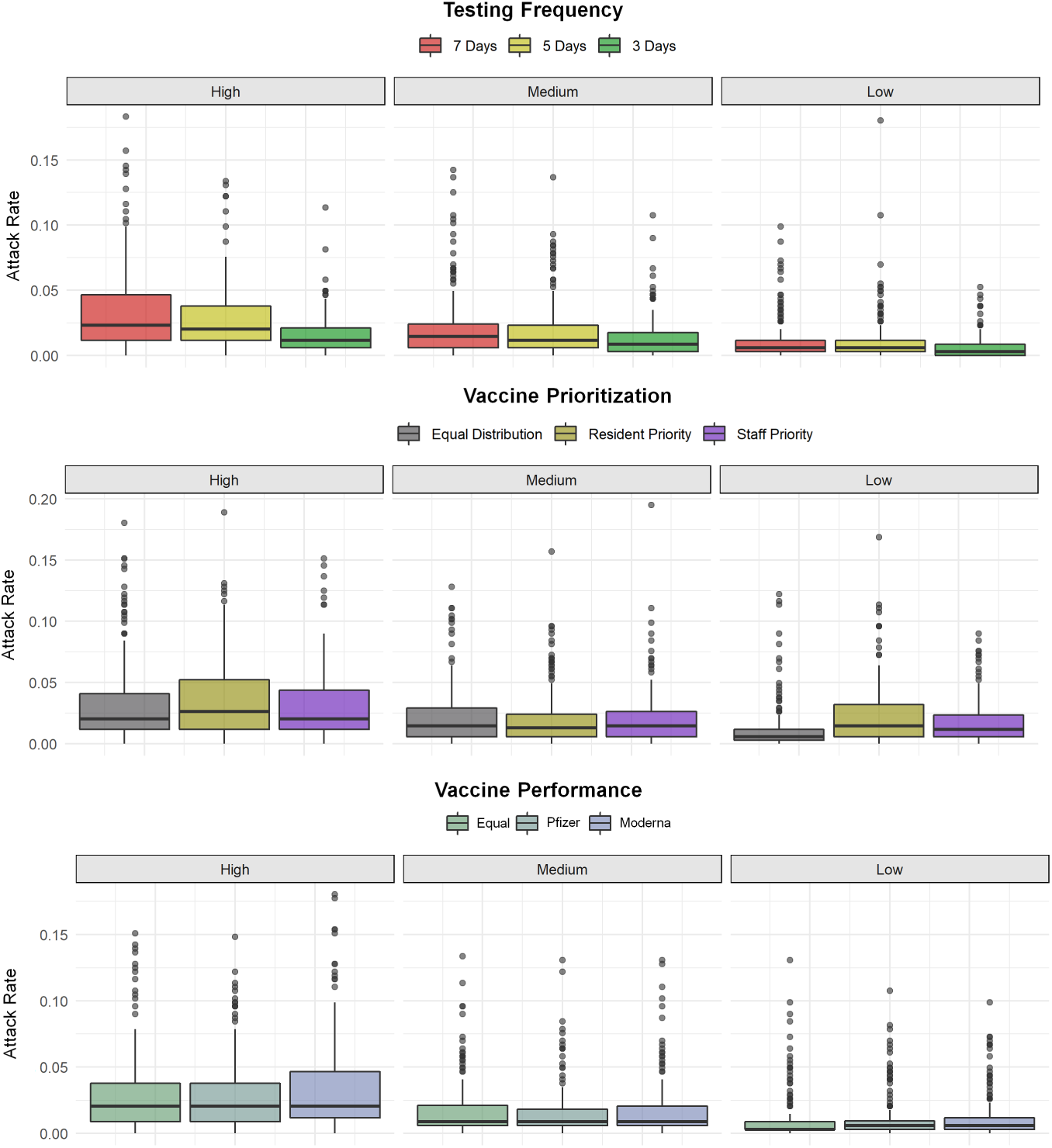
Attack rates for interventions under different assumptions of probability of introduction (low, medium, high).

### Testing and Vaccine Interventions

The implementation of frequent testing, particularly every 3-days reduced the attack rate by half and allowed containment of the outbreak within 9 days, despite high probability of virus introduction. Estimates and 95% confidence intervals illustrated in Figure 3 are provided in Supplemental Table 2. When vaccine was prioritized among staff, residents, or both, the attack did not seem to differ except when the introduction probability of was high, in which case the simulated median attack rate was 0.02 when staff were prioritized compared to 0.03 if residents were prioritized or no prioritization was present. Assuming a low probability of introduction, no prioritization provided the best opportunity to control an outbreak, leading to a median of 9 days (95% CI: 7-26) until eradication. We evaluate a vaccine’s ability to block transmission for scenarios of vaccine efficacy, staff and residents had the same efficacy, and residents had reduced efficacy compared to staff. We found that the probability of virus introduction was the most significant factor in determining the attack rate and days to the eradication of an outbreak. The attack rate doubled to 0.02 with high transmission probability and the time to the eradication of an outbreak was optimal only for low transmission. In all scenarios of low or moderate probability of transmission, none of the residents were infected.

**Figure 3:**
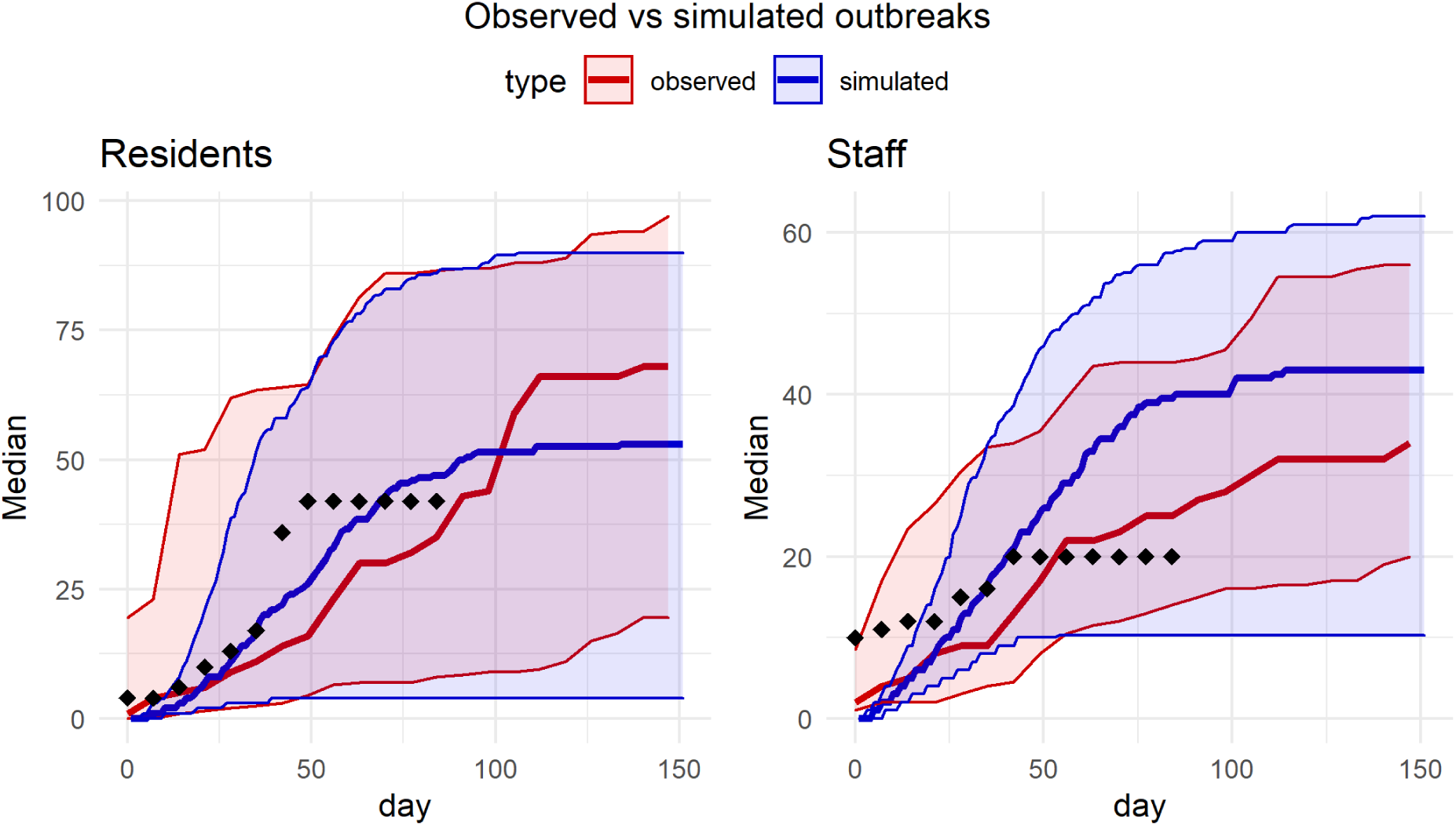
Model-predicted and observed number of cumulative incidence of confirmed cases for residents and staff. Dotted data represent the number of cases observed in the nursing home of study. Dark solid lines correspond to the median estimates for cases of staff and residents, and 25*th* and 75*th* percentiles are depicted in the shaded regions.

The model was well calibrated to the cumulative number of cases among residents and staff in California nursing homes, with *R*^2^ and Pearson’s *R* estimates higher than 0.79 (figure 3). Prospectively, our model overestimated confirmed cases among staff. likely due the implementation new interventions put in place after SARS-CoV-2 was introduced in a nursing home, namely increased frequency in testing. Our model underestimated cases among residents, and it is unclear if may be driven by the fact that some staff have more direct contacts with residents than others.

### Sensitivity Analysis

Baseline virus transmission rates, introduction probability, detection probability, PPE implementation and adherence, testing frequency, and vaccine efficacy were all considered for sensitivity analysis. We found that changes in the implementation of PPEs had a greater impact on reducing the attack rate and hospitalizations. Variation in virus transmission rates as well as the introduction probability showed substantial changes in attack rate and hospitalizations. Implementation of highly effective PPEs reduced the attack rate from 0.66 (95% CI: 0.39-0.85) to 0.02 (95% CI: 0-0.20), prevented 221 total infections, 18 hospitalizations, and reduced the period to eradicate the outbreak by almost a week. Increasing the probability of an introduction increased the total number of infections from 7 to 72, and 5 additional individuals were hospitalized. Analyses for these outcomes revealed significant decreases attributed to testing and vaccination across different frequency of testing and vaccine efficacy. Prevention of hospitalizations was more effectively accomplished through vaccination and was independent on age-specific vaccine efficacy assumptions.

## Discussion

The importance of careful use of non-pharmaceutical interventions was a critical lesson from the COVID-19 pandemic. Mask policies, limited visitation, and especially universal testing were critical to successful mitigation and prevention plans in the United States. Greater access to PPE and frequent testing surely played a part in reducing the case burden on LTCFs: case rates have dropped from a high of 33,625 nursing home cases/week to the current low of 1,927 cases/week [26]. December 18, 2020 marked the start of the Pharmacy Partnership for Long-Term Care Program in which the CDC partnered with multiple pharmacies to host on-site vaccination clinics for LTCF residents and staff [8]. Despite good vaccination progress, nursing home residents remain at high risk. As regulations ease, and with the possibility of requiring yearly vaccinations to prevent future outbreaks, we must consider how surveillance, PPE usage, and vaccine timing and prioritization complement each other. Our study sought to describe the potential combined effects of recommended NPIs and vaccine deployment strategies on the size and duration of a COVID-19 outbreak in a model nursing home.

Results from our model were most evident when we assumed a larger probability of viral introduction. In such cases, increased frequency of universal testing and isolation of positive cases (quarantine or paid leave) lead to larger reductions in attack rate than any other scenario. Prioritizing the vaccination of staff over residents lead to a moderate decrease in attack rate when viral introduction probability was high.

Community transmission rate is the strongest predictor of case rates in nursing homes thus far [27] and staff are the most important vectors through which introduction from the community occurs [11, 12, 28]. Our results support using strategic prioritization of staff for universal testing and vaccination as an important method for reducing the likelihood of an outbreak, especially in situations where community transmission is high.

There are several important challenges that these facilities will continue to face. LTCF administrators reported that staffing remains one of the primary barriers to maintaining high infection control standards [29]. Additionally, facilities that had a high degree of connectedness via shared staff showed higher case rates in general [30]. Expanded paid leave programs may also reduce the need for staff to seek additional employment to make ends meet, generally lowering their personal risk and the risk of introduction events.

Evidence indicates that staff may be more hesitant to get the vaccine than residents [10], and certainly have lower first-dose rates even if unrelated to hesitancy [8]. Vaccine mandates are one way to approach ensuring vaccine coverage goals are reached, but may create additional problems maintaining proper staff levels for delivering quality care. Additionally, nursing staff, including CNAs and LPNs, have high turnover rates in LTCFs. As a result, vaccination rates may fluctuate over time even within the same facility. Maintaining vaccine coverage goals will likely require an active program that includes acquiring accounting for staff who receive vaccines from a different source (i.e. a local pharmacy or a different job). We have even less data about the risks presented by reopening nursing homes to visitors, prompting questions about vaccine and testing requirements for visitors. An extension to this model that adds a visitor agent could help answer these questions before observational data becomes available.

### Strengths and Limitations

We calibrated our model to a real-world nursing home. The basal transmission model, in which no agents were vaccinated, generated plausible attack rates when compared to California nursing homes of a similar size. This, plus incorporating parameters from real-world data, provides external validity to the changes observed in our model. A particular strength of ABM is to show how complex outcomes can emerge from simple sets of rules; our model took advantage of this approach to show how interactions between staff and residents manifest the outbreak patterns observed in vivo. However, this model is primarily useful as an exploration of the impact of multiple interventions and introduction probabilities on an outbreak once introduction has occurred, and is therefore not meant to model the processes that lead to an introduction in the first place. Simulations were run for 150 days or until the facility was disease-free for up to 7 days; thus, it is also not able examine the impact of multiple introductions over longer periods of time or waning immunity from recovery or vaccination in its current form.

Not all data-derived parameters were made equally. The estimated effect of PPE use on transmission varied widely, thus making a reliable parameter difficult to define and the model sensitive to changes. Testing was also oversimplified in our model, as we assumed instantaneous results and all tests were equally sensitive. Additionally, we assumed that the effects of immunity, natural or from vaccination, was constant over the course of an outbreak and did not wane over time. We also assumed that staff agents had an equal chance of interacting with each resident agent, which is not reflective of intervention strategies that silo staff into daily routines focused on a specific subset of residents, such as dedicated staff for specific wards within the nursing home or for positive, isolated individuals.

## Data Availability

Code for reproducing this model is available in following repository: https://github.com/jpablo91/NH_COVID

## References

[1] T. N. Y. Times, “More than one-third of u.s. coronavirus deaths are linked to nursing homes.” https://www.nytimes.com/interactive/2020/us/coronavirus-nursing-homes.html. (Accessed on 03/09/2021).

[2] L. Harris-Kojetin, M. Sengupta, J. Lendon, V. Rome, and C. Caffrey, “Long-term care providers and services users in the united states, 2015–2016,” Vital Health Stat, 2019.

[3] B. F. Bigelow, O. Tang, B. Barshick, M. Peters, S. D. Sisson, K. S. Peairs, and M. J. Katz, “Outcomes of universal covid-19 testing following detection of incident cases in 11 long-term care facilities,” JAMA Intern Med, vol. 181, no. 1, pp. 127–129, 2020.

[4] J. K. Louie, H. M. Scott, A. DuBois, N. Sturtz, W. Lu, J. Stoltey, G. Masinde, S. Cohen, D. Sachdev, Philip N. Bobba, and S. F. D. o. P. H. C.-. S. N. F. O. R. T. Aragon, Tomas, “Lessons From Mass-Testing for Coronavirus Disease 2019 in Long-Term Care Facilities for the Elderly in San Francisco,” Clin. Infect. Dis., 07 2020. ciaa1020.

[5] J. G. Ouslander and D. C. Grabowski, “Covid-19 in nursing homes: Calming the perfect storm,” J Am Geriatr Soc, vol. 68, no. 10, pp. 2153–2162, 2020.

[6] C. Blackman, S. Farber, R. A. Feifer, V. Mor, and E. M. White, “An illustration of sars-cov-2 dissemination within a skilled nursing facility using heat maps,” J Am Geriatr Soc, vol. 68, no. 10, pp. 2174–2178, 2020.

[7] D. R. Smith, A. Duval, K. B. Pouwels, D. Guillemot, J. Fernandes, B.-T. Huynh, L. Temime, and L. Opatowski, “Optimizing covid-19 surveillance in long-term care facilities: a modelling study,” BMC Med., vol. 18, no. 386, 2020.

[8] R. Gharpure, A. Patel, and R. Link-Gelles, “First-dose covid-19 vaccination coverage among skilled nursing facility residents and staff,” JAMA, 2021.

[9] R. Gharpure, A. Guo, C. K. Bishnoi, U. Patel, D. Gifford, A. Tippins, A. Jaffe, E. Shulman, N. Stone, E. Mungai, S. Bagchi, J. Bell, A. Srinivasan, A. Patel, and R. Link-Gelles, “Early covid-19 first-dose vaccination coverage among residents and staff members of skilled nursing facilities participating in the pharmacy partnership for long-term care program - united states, december 2020-january 2021,” MMWR, vol. 70, pp. 178–182, 2021.

[10] K. T. Unroe, R. Evans, L. Weaver, D. Rusyniak, and J. Blackburn, “Willingness of long-term care staff to receive a covid-19 vaccine: A single state survey,” J Am Geriatr Soc, 2020.

[11] S. A. Goldberg, J. Lennerz, M. Klompas, E. Mark, V. M. Pierce, R. W. Thompson, et al., “Presymptomatic transmission of severe acute respiratory syndrome coronavirus 2 among residents and staff at a skilled nursing facility: Results of real-time polymerase chain reaction and serologic testing,” Clin. Infect. Dis., vol. 72, no. 4, p. 686–689, 2021.

[12] D. J. Toth and K. Khader, “Efficient sars-cov-2 surveillance strategies to prevent deadly outbreaks in vulnerable populations,” BMC Med., vol. 19, no. 25, 2021.

[13] M. Voysey, S. A. C. Clemens, S. A. Madhi, L. Y. Weckx, P. M. Folegatti, P. K. Aley, B. Angus, V. L. Baillie, S. L. Barnabas, Q. E. Bhorat, et al., “Safety and efficacy of the chadox1 ncov-19 vaccine azd1222) against sars-cov-2: an interim analysis of four randomised controlled trials in brazil, south africa, and the uk,” Lancet, vol. 397, no. 10269, pp. 99–111, 2021.

[14] C. for Disease Control and Prevention, “Cdc covid data tracker.” https://covid.cdc.gov/covid-data-tracker/#datatracker-home. (Accessed on 03/09/2021).

[15] X. He, E. H. Lau, P. Wu, X. Deng, J. Wang, X. Hao, Y. C. Lau, J. Y. Wong, Y. Guan, X. Tan, et al., “Temporal dynamics in viral shedding and transmissibility of covid-19,” Nat. Med., vol. 26, no. 5, pp. 672–675, 2020.

[16] M. Feaster and Y.-Y. Goh, “High proportion of asymptomatic sars-cov-2 infections in 9 long-term care facilities, pasadena, california, usa, april 2020,” Emerg Inf Dis, vol. 26, no. 10, p. 2416–2419, 2020.

[17] K. A. Walsh, K. Jordan, B. Clyne, D. Rohde, L. Drummond, P. Byrne, S. Ahern, P. G. Carty, K. K. O’Brien, E. O’Murchu, M. O’Neill, S. M. Smith, M. Ryan, and P. Harrington, “Sars-cov-2 detection, viral load and infectivity over the course of an infection,” Journal of Infection, vol. 81, no. 3, pp. 357–371, 2020.

[18] K. M. Azar, Z. Shen, R. J. Romanelli, S. H. Lockhart, K. Smits, S. Robinson, S. Brown, and A. R. Pressman, “Disparities in outcomes among covid-19 patients in a large health care system in california: Study estimates the covid-19 infection fatality rate at the us county level.,” Health Aff., vol. 39, no. 7, pp. 1253–1262, 2020.

[19] P. P. España, A. Bilbao, S. García-Gutiérrez, I. Lafuente, A. Anton-Ladislao, A. Villanueva, A. Uranga, M. J. Legarreta, U. Aguirre, and J. M. Quintana, “Predictors of mortality of covid-19 in the general population and nursing homes,” Intern Emerg Med, pp. 1–10, 2021.

[20] C. for Disease Control and Prevention, “Covid-19 pandemic planning scenarios — CDC.” https://www.cdc.gov/coronavirus/2019-ncov/hcp/planning-scenarios.html#table-2. (Accessed on 02/28/2021).

[21] M. Nuno, Y. Garcia, G. Rajasekar, D. Pinheiro, and A. J. Schmidt, “Covid-19 hospitalizations in five california hospitals,” medRxiv, 2021.

[22] D. K. Chu, E. A. Akl, S. Duda, K. Solo, S. Yaacoub, H. J. Schünemann, A. El-harakeh, A. Bognanni, T. Lotfi, M. Loeb, et al., “Physical distancing, face masks, and eye protection to prevent person-to-person transmission of sars-cov-2 and covid-19: a systematic review and meta-analysis,” Lancet, 2020.

[23] L. R. Baden, H. M. El Sahly, B. Essink, K. Kotloff, S. Frey, R. Novak, D. Diemert, S. A. Spector, N. Rouphael, C. B. Creech, et al., “Efficacy and safety of the mrna-1273 sars-cov-2 vaccine,” NEJM, 2020.

[24] F. P. Polack, S. J. Thomas, N. Kitchin, J. Absalon, A. Gurtman, S. Lockhart, J. L. Perez, G. Pérez Marc, E. D. Moreira, C. Zerbini, et al., “Safety and efficacy of the bnt162b2 mrna covid-19 vaccine,” NEJM, 2020.

[25] P. Taillandier, B. Gaudou, A. Grignard, Q. Huynh, N. Marilleau, P. Caillou, D. Philippon, and A. Drogoul, “Building, composing and experimenting complex spatial models with the gama platform,” Geoinformatica, vol. 23, no. 2, pp. 299–322, 2019.

[26] C. for Medicare Medicaid Services, “Covid-19 nursing home data — data.cms.gov.” https://data.cms.gov/stories/s/COVID-19-Nursing-Home-Data/bkwz-xpvg/. (Accessed on 03/09/2021).

[27] R. T. Konetzka and R. J. Gorges, “Nothing much has changed: Covid-19 nursing home cases and deaths follow fall surges,” J Am Geriatr Soc, 2020.

[28] D. J. Escobar, M. Lanzi, P. Saberi, R. Love, D. R. Linkin, J. J. Kelly, D. Jhala, V. Amorosa, M. Hofmann, and J. B. Doyon, “Mitigation of a coronavirus disease 2019 outbreak in a nursing home through serial testing of residents and staff,” Clin. Infect. Dis., vol. ciaa1021, 2020.

[29] G. K. SteelFisher, A. M. Epstein, D. C. Grabowski, K. E. Joynt Maddox, E. J. Orav, and M. L. Barnett, “Persistent challenges of covid-19 in skilled nursing facilities: The administrator perspective,” J Am Geriatr Soc, 2021.

[30] M. K. Chen, J. A. Chevalier, and E. F. Long, “Nursing home staff networks and covid-19,” PNAS, vol. 118, no. 1, p. e2015455118, 2021.

