## Supplementary information for "Testing and Vaccination to Reduce the Impact of COVID-19 in Nursing Homes: An Agent-Based Approach"

March 22, 2021

**Methods**

**Dynamic Transmission Model**
Resident and staff agents in our proposed model are represented in epidemiological classes of susceptible but not yet exposed to the disease ($S$), non-infectious exposed individuals incubating the disease whose infection is currently non-detectable by testing ($E$), infectious individuals with detectable disease who do not yet exhibit clinical symptoms of illness ($I_a$), infectious individuals exhibiting symptoms of illness ($I_s$), individuals that have recovered and can no longer infect others ($R$), symptomatic individuals requiring hospitalization ($H$), and individuals that succumbed to the disease ($D$) (Figure 1). Transition parameters are described in Table 1 of main manuscript.

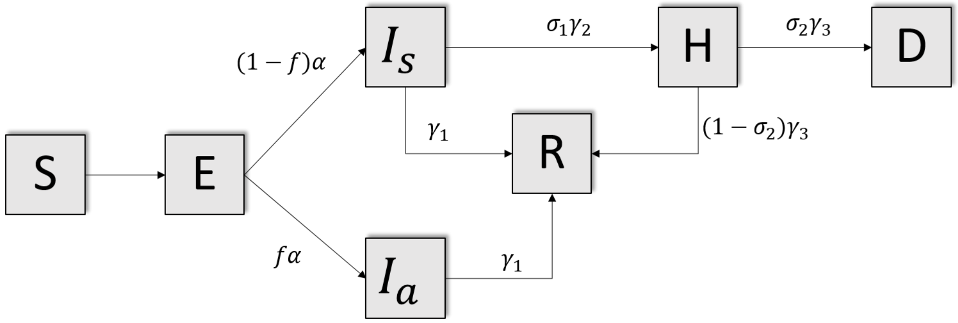


Figure 1: Epidemiological classes of the transmission model.

**Sensitivity Analysis**
The parameters used to describe the course of infection in our model were obtained from a combination of clinical trials and CDC reports. To describe the average time a person remains in the non-infectious latency state we used the infection profiles estimated by He et al. [1], which in our model was described by a log-normal distribution truncated between 2 to 10 days with shape 2, and scale 3. The probability of being asymptomatic in our model is based on data reported by the CDC [2] where a high proportion of asymptomatic SARS-CoV 2 (40.7%) where detected in 9 long term facilities. For parameters involved in the transmission probability such as shedding and infection we did not found enough information, therefore we calibrated these parameters in a way that the baseline scenario will produce outbreaks with similar sizes to the ones observed in nursery homes of similar number of residents.

To conduct the sensitivity analysis we set the parameters as the ones described on the table 1 from the main text, and selected one of the target parameters to vary as described on table 2. We ran the model 200 times and summarised the simulation results. The values for the PPE effect were based on the upper and lower bounds of the estimation of the PPE effect reported by the systematic review by Chu et al. [3]. For other parameters where no references were available, such as transmission, introduction and detection probability, we varied the parameter defining a worst and best case scenario.

**Scenarios**

Table 1 summarize the assumptions for the different scenarios. We analyze the impact of a variety of interventions under three rates of virus introduction which reflect the virus transmission in the community. The first scenario considers different testing periods, the second regard different vaccine efficacy, and the last one contemplates different prioritization strategies in vaccination.

Table 1: Description of interventions and parameter values.

| Probability of Introduction | Low | Medium | High |
|---|---|---|---|
| Testing Frequency ($\omega$ = Introduction probability) | | | |
| | $\omega = 0.05$ | $\omega = 0.10$ | $\omega = 0.15$ |
| Testing Interval | 7 days | 7 days | 7 days |
| Testing Interval | 5 days | 5 days | 5 days |
| Testing Interval | 3 days | 3 days | 3 days |
| Vaccine Performance ($V_rOR$ = Odds ratio for the vaccine effect on residents, $V_sOR$= Odds ratios for the vaccine effect on staff) | | | |
| | $\omega = 0.05$ | $\omega = 0.10$ | $\omega = 0.15$ |
| Equal | $V_rOR = 0.0493$<br>$V_sOR = 0.0493$ | $V_rOR = 0.0493$<br>$V_sOR = 0.0493$ | $V_rOR = 0.0493$<br>$V_sOR = 0.0493$ |
| Pfizer | $V_rOR = 0.0619$<br>$V_sOR = 0.0434$ | $V_rOR = 0.0619$<br>$V_sOR = 0.0434$ | $V_rOR = 0.0619$<br>$V_sOR = 0.0434$ |
| Moderna | $V_rOR = 0.1357$<br>$V_sOR = 0.0441$ | $V_rOR = 0.1357$<br>$V_sOR = 0.0441$ | $V_rOR = 0.1357$<br>$V_sOR = 0.0441$ |
| Vaccine Prioritization ($V_r\%$=Percentage of residents vaccinated, $V_s\%$=Percentage of staff vaccinated) | | | |
| | $\omega = 0.05$ | $\omega = 0.10$ | $\omega = 0.15$ |
| Equal | $V_r\% = 50\%$<br>$V_s\% = 50\%$ | $V_r\% = 50\%$<br>$V_s\% = 50\%$ | $V_r\% = 50\%$<br>$V_s\% = 50\%$ |
| Staff Priority | $V_r\% = 30\%$<br>$V_s\% = 70\%$ | $V_r\% = 30\%$<br>$V_s\% = 70\%$ | $V_r\% = 30\%$<br>$V_s\% = 70\%$ |
| Resident priority | $V_r\% = 70\%$<br>$V_s\% = 30\%$ | $V_r\% = 70\%$<br>$V_s\% = 30\%$ | $V_r\% = 70\%$<br>$V_s\% = 30\%$ |

Table 2: Results from scenario modeling using the median and 95% confidence intervals.

| Scenario | Days to eradication | Attack Rate | Total Infected | Infected residents | Infected staff | Hospitalizations | Deaths |
|---|---|---|---|---|---|---|---|
| Introduction Probability & Testing Frequency | | | | | | | |
| High & 7-days | 14 (7, 48) | 0.02 (0, 0.1) | 8 (1, 34) | 2 (0, 16) | 6 (1, 21) | 0.5 (0, 3) | 0 (0, 1) |
| High & 5-days | 15 (5, 41) | 0.02 (0, 0.07) | 7 (1, 25) | 1 (0, 12) | 5 (1, 18) | 0 (0, 3) | 0 (0, 1) |
| High & 3-days | 9 (3, 24) | 0.01 (0, 0.04) | 4 (1, 14) | 0 (0, 6) | 3 (1, 11) | 0 (0, 2) | 0 (0, 0) |
| Moderate & 7-days | 14 (7, 41) | 0.01 (0, 0.07) | 5 (1, 23) | 1 (0, 11) | 3 (1, 14) | 0 (0, 2) | 0 (0, 0) |
| Moderate & 5-days | 10 (5, 35) | 0.01 (0, 0.07) | 4 (0, 24) | 0 (0, 12) | 3 (0, 13) | 0 (0, 2) | 0 (0, 0) |
| Moderate & 3-days | 9 (3, 30) | 0.01 (0, 0.04) | 3 (0, 15) | 0 (0, 6) | 2 (0, 10) | 0 (0, 1) | 0 (0, 0) |
| Low & 7-days | 9 (7, 27) | 0.01 (0, 0.04) | 2 (0, 14) | 0 (0, 9) | 1 (0, 7) | 0 (0, 2) | 0 (0, 0) |
| Low & 5-days | 9 (5, 25) | 0.01 (0, 0.04) | 2 (0, 13) | 0 (0, 8) | 1 (0, 7) | 0 (0, 2) | 0 (0, 0) |
| Low & 3-days | 9 (3, 20) | 0 (0, 0.02) | 1 (0, 8) | 0 (0, 4) | 1 (0, 5) | 0 (0, 1) | 0 (0, 0) |
| Introduction Probability & Vaccine Prioritization | | | | | | | |
| High & Equal distribution | 14 (7, 42) | 0.02 (0, 0.11) | 7 (1, 39) | 1 (0, 17) | 6 (1, 22) | 0 (0, 3) | 0 (0, 1) |
| High & Resident | 14 (7, 56) | 0.03 (0, 0.1) | 9 (1, 35) | 2 (0, 14) | 7 (1, 22) | 0 (0, 3) | 0 (0, 1) |
| High & Staff | 14 (7, 49) | 0.02 (0, 0.09) | 7 (1, 30) | 1 (0, 15) | 5 (1, 18) | 0 (0, 3) | 0 (0, 0) |
| Moderate & Equal distribution | 14 (7, 42) | 0.01 (0, 0.07) | 5 (1, 24) | 1 (0, 12) | 3 (1, 15) | 0 (0, 3) | 0 (0, 0.05) |
| Moderate & Resident | 14 (7, 35) | 0.01 (0, 0.07) | 5 (1, 24) | 1 (0, 12) | 4 (1, 14) | 0 (0, 3) | 0 (0, 1) |
| Moderate & Staff | 14 (7, 35) | 0.01 (0, 0.06) | 5 (1, 22) | 1 (0, 11) | 3 (0, 11) | 0 (0, 2) | 0 (0, 0.05) |
| Low & Equal distribution | 9 (7, 26) | 0.01 (0, 0.04) | 2 (0, 15) | 0 (0, 9) | 1 (0, 8) | 0 (0,2) | 0 (0, 0) |
| Low & Resident | 14 (7, 42) | 0.01 (0, 0.07) | 5 (1, 25) | 1 (0, 11) | 4 (0, 15) | 0 (0, 3) | 0 (0, 1) |
| Low & Staff | 14 (7, 35) | 0.01 (0, 0.06) | 4 (1, 19) | 1 (0, 10) | 3 (0, 10) | 0 (0, 2) | 0 (0, 0) |
| Introduction Probability & Vaccine Performance | | | | | | | |
| High & Similar efficacy | 14 (7, 42) | 0.02 (0, 0.1) | 7 (1, 35) | 1 (0, 15) | 5 (1, 20) | 0 (0, 3) | 0 (0, 0) |
| High & Reduced efficacy (Pfizer) | 14 (7, 42) | 0.02 (0, 0.1) | 7 (1, 33) | 1 (0, 15) | 6 (1, 18) | 0 (0, 4) | 0 (0, 0) |
| High & Reduced efficacy (Moderna) | 14 (7, 45) | 0.02 (0, 0.12) | 7 (1, 40) | 1 (0, 19) | 6 (1, 21) | 0 (0, 4) | 0 (0, 1) |
| Moderate & Similar efficacy | 14 (7, 36) | 0.01 (0, 0.06) | 3 (1, 20) | 0 (0, 8) | 3 (0, 13) | 0 (0, 2) | 0 (0, 0) |
| Moderate & Reduced efficacy (Pfizer) | 13 (7, 35) | 0.01 (0, 0.06) | 3 (1, 20) | 0 (0, 6) | 3 (0, 11) | 0 (0, 2) | 0 (0, 0) |
| Moderate & Reduced efficacy (Moderna) | 14 (7, 36) | 0.01 (0, 0.06) | 3 (1, 21) | 0 (0, 9) | 3 (0, 14) | 0 (0, 2) | 0 (0, 0) |
| Low & Similar efficacy | 9 (7, 27) | 0 (0, 0.05) | 1 (0, 17) | 0 (0, 10) | 1 (0, 8) | 0 (0, 2) | 0 (0, 0) |
| Low & Reduced efficacy (Pfizer) | 9 (7, 28) | 0.01 (0, 0.05) | 2 (0, 16) | 0 (0, 9) | 1 (0, 7) | 0 (0, 2) | 0 (0, 0) |
| Low & Reduced efficacy (Moderna) | 9 (7, 28) | 0.01 (0, 0.05) | 2 (0, 16) | 0 (0, 10) | 1 (0, 8) | 0 (0, 2) | 0 (0, 0) |

### References


[1] X. He, E. H. Lau, P. Wu, X. Deng, J. Wang, X. Hao, Y. C. Lau, J. Y. Wong, Y. Guan, X. Tan, *et al.*, "Temporal dynamics in viral shedding and transmissibility of covid-19," *Nat. Med.*, vol. 26, no. 5, pp. 672–675, 2020.

[2] M. Feaster and Y.-Y. Goh, "High proportion of asymptomatic sars-cov-2 infections in 9 long-term care facilities, pasadena, california, usa, april 2020," *Emerg Inf Dis*, vol. 26, no. 10, p. 2416–2419, 2020.

[3] D. K. Chu, E. A. Akl, S. Duda, K. Solo, S. Yaacoub, H. J. Schünemann, A. El-harakeh, A. Bognanni, T. Lotfi, M. Loeb, *et al.*, "Physical distancing, face masks, and eye protection to prevent person-to-person transmission of sars-cov-2 and covid-19: a systematic review and meta-analysis," *Lancet*, 2020.
